# Clinical characteristics of Covid-19 patients with re-positive test results: an observational study

**DOI:** 10.1101/2020.06.23.20138149

**Authors:** Ying Su, Ling-Shuang Zhu, Yong Gao, Yuncheng Li, Zhanlu Xiong, Bo Hu, Jiao Yuan, Zhao Zhang, Xiaoxiong Wu, Chunchen Feng, Fanjun Cheng, Yang Jin, Yu Zhang, Youming Lu, Hongbo Wang, Ling-Qiang Zhu

## Abstract

**Background:** With coronavirus disease 2019 (Covid-19) ravaging the global, concern has been aroused whether discharged Covid-19 patients with reappeared positive nucleic acid test results are infected again.

**Objective:** To analyze the clinical characteristics of discharged Covid-19 patients with reappeared positive nucleic acid test results and to track clinical outcomes of them.

**Methods:** We extracted clinical data on 938 Covid-19 patients from Wuhan Union Hospital (West Branch), and we obtained information about residual symptoms and nucleic acid tests after discharge through follow-up study. We evaluated the relationship of clinical characteristics and reappeared positive results. Each patient had at least 44 days of follow-up.

**Results:** Of 938 discharged patients, a total of 58 (6.2%) had reappeared positive nucleic acid test results and 880 remain negative. Among patients over the age of 50, the factors we found to be associated with re-positive results were coronary artery disease (14.1%, vs. 5.5% among those without coronary artery disease; odds ratio, 2.81; 95% confidence interval [CI], 1.28 to 6.15), and hypertension (9.5%, vs. 4.9% among those without hypertension; odds ratio, 2.05; 95% CI, 1.10 to 3.82). As of May 11, 2020, 54 (93.1%) re-positive patients turned negative again while two patients remained positive, and two patients was lost to the second follow-up.

**Conclusion:** Coexisting diseases including coronary artery disease and hypertension were substantial risk factors for re-positive outcomes among patients over 50. And most re-positive patients tended to return negative eventually.

## Introduction

Covid-19 pandemic continues to ravage the global, with 8974795 confirmed cases and 469159 documented deaths as of June 23, 2020 (1). Real-time reverse transcription polymerase chain reaction (RT-PCR) test for severe acute respiratory syndrome coronavirus 2 (SARS-CoV-2) is widely carried out as a diagnostic and discharge criterion (2). However, recent reports regarding positive SARS-CoV-2 test results in some recovered cases after discharged home were published (3-6). Concern has been aroused that these recovered patients with a reappeared positive test might be infected and contagious again, given that no evidence suggests people with Covid-19 antibodies protected from second infection (7).

Among the main possibilities on the re-detectable positive results are second infection, relapse, or RNA fragment detection. Second infection would be the worst-case scenario due to the impediment to vaccine development and herd immunity (8, 9). It has been suggested that patients tested positive again were unlikely to relapse, because no live virus was isolated in them and positive tests may result from inactive virus fragment. But a recent report suggests that patients may still carry SARS-CoV-2 deep in the lungs after recovery and discharge, and the presence of virus cannot be detected by conventional nucleic acid test from nasopharyngeal swabs (10).

It is crucial to figure out susceptibility of convalescent patients to the re-positive experience as well as clinical outcomes of it. Therefore, we undertook a research to study the relationship between reappeared positive test results and clinical features.

## Methods

### Study oversight

The study was approved by the institutional review board of Wuhan Union Hospital and designed by the authors. Data were analyzed and interpreted by the authors. All the authors had full access to the data and vouch for the accuracy and completeness of the data. All the author claimed responsible for the decision to submit the manuscript for publication.

### Study population and data collection

We collected the medical records and compiled data of 1208 hospitalized patients with laboratory-confirmed Covid-19 in Wuhan Union Hospital (West Branch), who were discharged between January 18, 2020 and March 26, 2020. A positive Covid-19 case was defined as a positive result on high-throughput sequencing or RT-PCR assay of nasal and pharyngeal swab specimens. As of April 24, 2020, we carried out the first telephone follow-up survey on these 1208 discharged patients, 938 cases of which finished follow-up study. 270 cases were lost mainly due to wrong phone numbers, unanswered calls, and incoordinate patients. A second follow-up was targeted on 58 re-positive patients as of May 11, 2020, and 56 patients finished it.

We extracted patients’ demographic and clinical characteristics from electronic medical records, including age, sex, length of hospital stays, severity on admission, and coexisting diseases. Coexisting diseases included cardiovascular disease (including cardiac dysfunction, coronary artery disease, and hypertension), respiratory disease (asthma and chronic bronchitis), autoimmune disease, cancer, diabetes mellitus, heptic dysfunction, and renal dysfunction. We obtained clinical information after discharge through follow-up survey, including residual symptoms and laboratory re-examinations results on SARS-COV-2 nucleic acid, chest imaging, and serum antibody. Residual symptoms included chest congestion, palpitation, expectoration, cough, fatigue, dry eyes, myalgia, sore throat, headache, diarrhea, and fever.

### Study definitions

Fever was defined as an axillary temperature of 37·3°C or higher.

Mild and moderate Covid-19 patients were defined as patients who had mild clinical symptoms with or without pneumonia on chest imaging. Severe and critically ill patients were patients meeting up to any of the following standards: respiratory rate (RR)≥30 times/minute; finger oxygen saturation≤93% at rest; the ratio of the partial pressure of arterial oxygen to the fraction of inspired oxygen [Pao2:Fio2], ≤ 300mmHg (1mmHg=0·133kPa).

Discharge criterion was defined as follows: Temperature returned back to normal for over three days with respiratory symptoms significantly improved; chest imaging showed inflammation absorption; and two consecutive nucleic acid tests for SARS-COV-2, sampling interval of which exceeded 24 hours, were negative.

### Statistical analysis

Continuous variables were reported as means and standard deviations. Categorical variables were summarized as counts and percentages.

Independent-sample T test or T’ test was completed to assess the differences between male and female groups. Mann-Whitney U test, χ2 test, or Fisher’s exact test were used to compare differences between re-positive and negative groups where appropriate. The category of analyzed diseases and symptoms was decided according to re-positive patients. If the patient’s electronic medical record and follow-up record did not include information on a clinical characteristic, such as hypertension or cough after discharge, that characteristic was assumed not present.

Age- and sex-adjusted linear regression analyses were performed to confirm the relationship between re-positive test results and symptoms after discharge (chest congestion, palpitation, expectoration, cough, fatigue, dry eyes, myalgia, sore throat, headache, diarrhea, and fever). Age- and sex-adjusted linear regression analyses were performed to confirm the relationship between coexisting diseases (diabetes mellitus, coronary artery disease, hypertension, and coexisting conditions) and re-positive tests, with odds ratios and 95% confidence intervals calculated. The 95% confidence intervals have not been adjusted for multiple testing and should not be used to infer definitive effects.

All statistical analyses were performed with SPSS Statistics software, version 26 (IBM).

## Results

### 1. Demographic and Clinical Characteristics

As of April 24, 2020, our first follow-up study population included 938 patients (510 female cases) hospitalized for Covid-19 in Wuhan Union Hospital (West Branch) who were discharged between January 18, 2020 and March 26, 2020 (Figure 1). Among them, 14.9% were mild and moderate patients, 85.1% were severe and critically ill (Table 1). The mean (±SD) age of the patients was 57.0±14.6 years (range, 16 to 94 years). A majority of the patients (72.6%) were above 50 years. Severe and critically ill patients were older than mild and moderate patients by 5.3 years. And the mean (±SD) length of hospital stay was 21.3±11.5 days (range, 1 to 59 days) (Table S1-2).

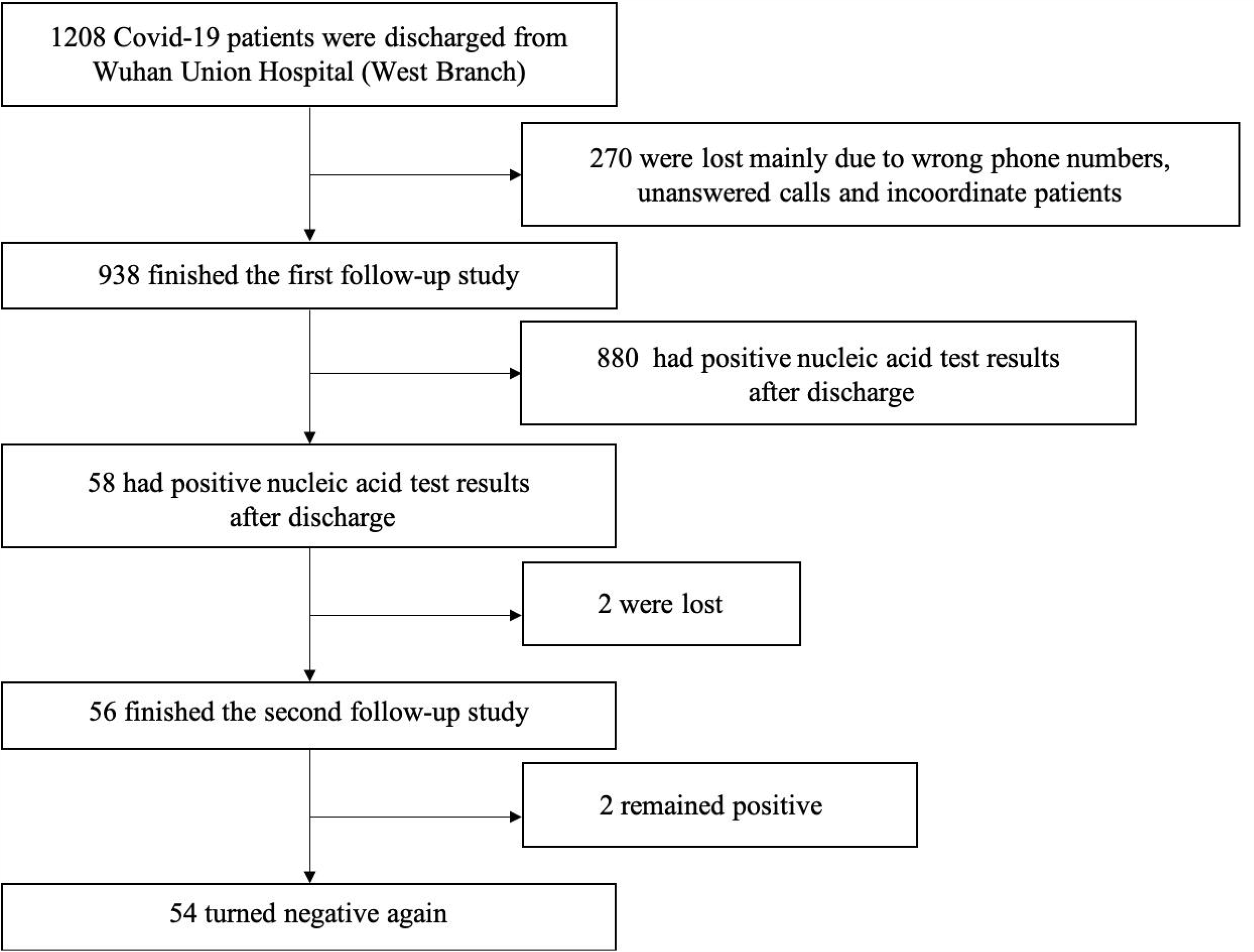

**Table 1.** Demographic and Clinical Characteristics of 938 Patients

| Characteristic | All Patient | Male | Female |
| --- | --- | --- | --- |

|  | (N=938) | (n=428) | (n=510) |
| --- | --- | --- | --- |
| Severity of illness, n (%) |  |  |  |
| Mild and moderate | 140 (14.9) | 54 (12.6) | 86 (16.9) |
| Severe and critically ill | 798 (85.1) | 374 (87.4) | 424 (83.1) |
| Mean age (range), year | 57.0±14.6 (16-94) | 56.9±14.0 (16-94) | 57.1±15.1 (18-94) |
| Mean length of hospital stay (range), day | 21.3±11.5 (1-59) | 21.2±11.2 (4-59) | 21.3±11.7 (1-54) |
| Positive retests after discharge, n (%) | 58 (6.2) | 30 (7.0) | 28 (5.5) |
| Residual symptoms after discharge, n (%) |  |  |  |
| Chest congestion | 192 (20.5) | 68 (15.9) | 124 (24.3) |
| Cough | 124 (13.2) | 43 (10.0) | 81 (15.9) |
| Diarrhea | 34 (3.6) | 18 (4.2) | 16 (3.1) |
| Dry eyes | 109 (11.6) | 42 (9.8) | 67 (13.1) |
| Expectoration | 128 (13.6) | 50 (11.7) | 78 (15.3) |
| Fatigue | 111 (11.8) | 35 (8.2) | 76 (14.9) |
| Fever | 16 (1.7) | 7 (1.6) | 9 (1.8) |
| Headache | 53 (5.7) | 11 (2.6) | 42 (8.2) |
| Myalgia | 72 (7.7) | 21 (4.9) | 51 (10.0) |
| Palpitation | 154 (16.4) | 50 (11.7) | 104 (20.4) |
| Sore throat | 61 (6.5) | 24 (5.6) | 37 (7.3) |

Plus-minus values are means ±SD.

462 (49.3%) patients had residual symptoms after discharge. The most common symptoms were chest congestion and palpitation, which respectively occurred in 192 (20.5%) and 154 (16.4%) patients. 128 (13.6%) had expectoration, 124 (13.2%) patients had cough, 111 (11.8%) patients had fatigue, and 109 (11.6%) had dry eyes. Other symptoms included myalgia (7.7%), sore throat (6.5%), headache (5.7%), diarrhea (3.6%), and fever (1.7%) (Table 1). Women had a great prevalence of chest congestion, palpitation, cough, fatigue, myalgia, and headache, whereas no association was found between gender and other five symptoms (Table S2).

### 2. Characteristics of Retested Positive (RP) Patients Compared with Retested Negative (RN) Patients

Fifty-eight (6.2%) patients were tested positive again in nucleic acid tests after discharge. The mean (±SD) age and mean (±SD) length of hospital stay of RP patients was respectively 57.8±13.4 years (range, 19 to 79 years) and 19.2±9.5 years (range, 4 to 42 days), with no difference between RP and RN group (Table S3).

Coexisting diseases were common among both RP and RN patients. Twenty-one (36.2%) RP patients had hypertension and 11 (19.0%) had diabetes mellitus. Nine (15.5%) had coronary artery disease, three (5.2%) had cardiac dysfunction, two (3.4%) had cancer, and one (1.7%) had chronic bronchitis. And no association between coexisting diseases and RP patients was found within the full sample. Residual symptoms after discharge were more common in RP patients. Twenty-three (39.7%) RP patients had chest congestion, 15 (25.9%) had cough, 13 (22.4%) had dry eyes, 13 (22.4%) had palpitation, 11 (19.0%) had fatigue, and 11 (19.0%) had expectoration. Particularly, RP patients were more likely to regain chest congestion, cough, diarrhea, dry eyes, and fever after discharge (Table S3-4).

### 3. Linear Regression Analysis Based on Re-Positive Patients over 50

The most common coexisting diseases among RP patients were hypertension, diabetes mellitus, and coronary artery disease. Noticeably, all of these diseases were mainly distributed in RP patients over 50 years old. And RP patients had higher prevalence than RN patients, especially coronary artery disease and hypertension (Table S5).

Age- and sex-adjusted linear regression models were used to analyze relationship between coexisting diseases and re-positive nucleic acid results. The odds ratios and coefficients were present in Table 2 and Table S6. Coronary artery disease and hypertension were associated with a higher risk of re-positive test results. For coronary artery disease, the odds ratio was 2.81 (95% confidence interval [CI], 1.28 to 6.15) and coefficient was 0.089 (95% CI, 0.026 to 0.151). For hypertension, the odds ratio was 2.05 (95% confidence interval [CI], 1.10 to 3.82) and coefficient was 0.048 (95% CI, 0.008 to 0.088). Although no association was found in diabetes mellitus (odds ratio, 1.57; 95% CI, 0.75 to 3.29), we were surprised to find that coexisting of diabetes mellitus rose the odds ratio and coefficient of coronary artery disease to 8.46 (95% CI, 3.01 to 23.80) and 0.286 (95% CI, 0.173 to 0.398) respectively.

**Table 2.**
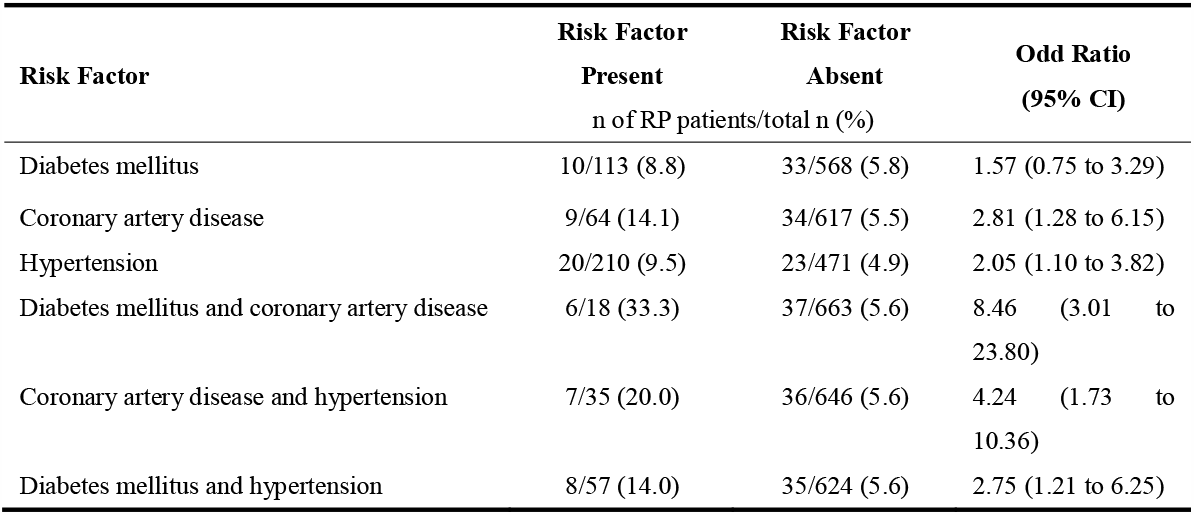
Independent Predictors of Re-Positive Result from Linear Regression

**Table 3.** Characteristics of RP Patients Who Turned Negative again

| Characterisitic | RP Patients Turning Negative |
| --- | --- |
| Chest CT imaging after discharge, n/total n (%) |  |
| Improvement | 37/41 (90.2) |
| No change | 4/41 (9.7) |
| Deterioration | 0/41 (0) |
| Serum IgG and IgM after discharge, n/total n (%) |  |
| IgG (+) IgM (-) | 22/29 (75.9) |
| IgG (-) IgM (-) | 2/29 (6.9) |
| IgG (+) IgM (+) | 5/29 (17.2) |
| Mean interval of turning positive after discharge (range), day | 12.2±5.7 (3-39) |
| Mean interval of turning negative again (range), day | 12.9±13.2 (1-53) |
| Residual symptoms after turning negative, n/total n (%) |  |
| Chest congestion | 2/54 (3.7) |
| Cough | 2/54 (3.7) |
| Expectoration | 1/54 (1.9) |
| Fatigue | 3/54 (5.5) |
| Palpitation | 1/54 (1.9) |
| Sore throat | 1/54 (1.9) |

Plus-minus values are means ±SD.

### 4. Clinical Outcomes of 58 RP Patients

Forty-one RP patients received chest CT imaging after discharge, with 37 (90.2%) cases showing improvement. And 29 patients received serum IgG and IgM tests. Twenty-two (75.9%) patients were IgG positive and IgM negative, five (17.2%) patients were both positive, while two (6.9%) patients were both negative.

Two patients were lost to the second follow-up for RP patients. Among the rest patients, 54 (93.1%) turned negative again while two patients remained positive as of May 11, 2020. The mean interval (±SD) of turning positive after discharge was 12.2±5.7 days (range, 3 to 39 days) and mean interval (±SD) of turning negative again was 12.9±13.2 days (range, 1 to 53 days). As of May 11, 2020, the percentage of RP patients with residual symptoms sharply declined from 67.0% at discharge to 16.7%. Three (5.5%) patients had fatigue, two (3.7%) had cough, two (3.7%) had chest congestion. There was only one (1.9%) patient with expectoration, one (1.9%) with throat, and one (1.9%) with palpitation.

## Discussion

In this follow-up analysis that included 938 Covid-19 patients, no evidence indicated that age, sex or severity was associated with re-positive outcomes. Harmful associations were noted for coronary artery disease and hypertension among patients over 50 years old. Considering the urgency of the pandemic and the uncertainty of re-positive patients, we chose to report our findings before collecting a larger sample to provide clinical advice to focus on patients with coronary heart disease and hypertension over the age of 50.

Comorbidities such as coronary heart diseases, hypertension and diabetes predisposed to detrimental clinical outcomes in Covid-19 patients (11-13). Evidence was found that patients may still carry SARS-CoV-2 deep in the lungs after recovery, which cannot be detected by conventional nucleic acid test for upper respiratory tract (12). Some patients may still carry a low load of SARS-Cov-2 virus at discharge, whereas impaired immunity due to comorbidities could result in a larger fluctuation in virus load and therefore a re-positive state several days later. Besides, a recent study pointed out that hypertension was a prominent risk factor preventing viral clearance in Covid-19 patients (14). SARS-Cov-2 enters cells using angiotensin-converting enzyme2 (ACE2) (15, 16), which is a widely-expressed membrane-bound aminopeptidase in heart, kidney and lung (17, 18). Treatment of ACE inhibitors or angiotensin II type-1 receptor blockers (ARBs) on patients with hypertension results in a substantially increased expression of ACE2 (19, 20), which increases risk of virus invasion and difficulty of clearance.

Despite that mechanisms underlying re-positive outcome remain unclear, the possibility of secondary infection is small because these RP patients caused no infection after discharge and most of them returned negative again with an alleviation in symptoms. As of May 11, 2020, the percentage of RP patients with residual symptoms sharply declined, and 93.1% RP patients turned negative again.

Our study has several limitations. First, the short duration of follow-up makes it hard to determine clinical ends of RP patients. It is uncertain whether they will remain negative or turn positive again. Long-term follow-up are needed to evaluate the possible risk of re-positive patients. Second, some patients had incomplete medical documentation. The anamnesis was considered inexistent if not documented. Third, because Wuhan Union Hospital (West Branch) mainly treats severe and critically ill patients, only 140 mild and moderate patients were included in the follow-up study. It is uncertain whether the conclusions can be generalized to mild and moderate patients. Finally, due to our focus on characteristics of re-positive patients, more attention was paid to harmful factors and some protective factors may be missed.

In summary, we demonstrated coronary artery disease and hypertension were associated with an increased risk of re-positive test results for patients over 50. However, re-positive patients tended to turn negative eventually. Our findings also highlight the importance of following up discharged patients because two consecutive negative nucleic acid tests at discharge were not necessarily the endpoints.

## Data Availability

After publication, the data supporting the findings of this study can be available on reasonable requests to the corresponding authors. A proposal with detailed description of study objectives and statistical analysis plan will be needed for evaluation of the reasonability of requests. Additional materials may also be required. The corresponding authors and Wuhan Union Hospital will make a decision based on these materials. An email address will be provided for communication if the data are approved to be shared with others. We can provide data without names and identifiers. Study protocol, statistical analysis plan, and informed consent form are unavailable. The corresponding authors have the right to decide whether to share the data or not based on the research objectives and plan provided.

## Acknowledgments

This study is supported partially by the National Natural Science Foundation of China (81871108, 81829002, 81961128005, 81761138043, 31721002), Top-Notch Young Talents Program of China of 2014, and Academic Frontier Youth Team of Huazhong University of Science and Technology to Dr. Ling-Qiang Zhu.

---

**Supplementary Table 1.** Age and Hospital Stay Data Based on Severity

| Characteristic | Mild and Moderate Patients | Severe and Critically Ill Patients |
| --- | --- | --- |
| Age |  |  |
| Mean (range) -- year | 52.5±14.2 (19-86) | 57.8±14.6 (16-94) |
| Distribution -- n (%) |  |  |
| 0-24 | 3 (2.1) | 7 (0.9) |
| 25-49 | 51 (36.4) | 196 (24.6) |
| 50-74 | 82 (58.6) | 507 (63.5) |
| ≥75 | 4 (2.9) | 88 (11.0) |
| Mean length of hospital stay (range), day | 14.9±8.3 (1-44) | 22.4±11.6 (2-59) |

Plus-minus values are means ±SD.

**Table S2.** Linear Regression Analysis Based on 938 Patients

a) Linear regression model for severity
| Variable | Coefficients | 95% CI (Low) | 95% CI (High) | p Value |
| --- | --- | --- | --- | --- |
| Sex (male as reference) | 0.042 | -0.003 | 0.088 | 0.069 |
| Age | 0.003 | 0.002 | 0.005 | <0.001 *** |

b) Linear regression model of 11 residual symptoms on sex (male as reference)
| Residual symptoms | Coefficients | 95% CI (Low) | 95% CI (High) | p Value |
| --- | --- | --- | --- | --- |
| Chest congestion | -0.084 | -0.136 | -0.033 | 0.001 ** |
| Cough | -0.058 | -0.102 | -0.015 | 0.009 ** |
| Diarrhea | 0.011 | -0.013 | 0.035 | 0.384 |
| Dry eyes | -0.033 | -0.074 | 0.008 | 0.114 |
| Expectoration | -0.036 | -0.080 | 0.008 | 0.109 |
| Fatigue | -0.067 | -0.109 | -0.026 | 0.001 ** |
| Fever | -0.001 | -0.018 | 0.015 | 0.879 |
| Headache | -0.057 | -0.086 | -0.027 | <0.001 *** |
| Myalgia | -0.051 | -0.085 | -0.017 | 0.003 ** |
| Palpitation | -0.087 | -0.134 | -0.040 | <0.001 *** |
| Sore throat | -0.016 | -0.048 | 0.015 | 0.309 |

\* p < 0.05 \*\*p < 0.01 \*\*\*p < 0.001

**Supplementary Table 3.** Characteristic of RP and RN patients

| Characteristic | RP Patients<br>(n=58) | RN Patients<br>(n=880) | Difference<br>(95% CI) |
| --- | --- | --- | --- |
| Mean age (range), year | 57.8±13.4 (19-79) | 56.9±14.7 (16-94) | 0.9 (-3.0 to 4.8) |
| Mean length of hospital stay (range), day | 19.2±9.5 (4-42) | 21.4±11.6 (1-59) | -2.2 (-5.3 to 0.8) |
| Coexisting diseases, n (%) |  |  |  |
| Cancer | 2 (3.4) | 24 (2.7) | 0.7 (-3.7 to 5.1) |
| Cardiac dysfunction | 3 (5.2) | 12 (1.4) | 3.8 (-2.1 to 9.7) |
| Chronic bronchitis | 1 (1.7) | 10 (1.1) | 0.6 (-2.9 to 4.1) |
| Coronary artery disease | 9 (15.5) | 107 (12.2) | 3.3 (-5.4 to 12.1) |
| Diabetes mellitus | 11 (19.0) | 127 (14.7) | 4.8 (-5.2 to 13.8) |
| Hypertension | 21 (36.2) | 246 (28.0) | 8.2 (-4.8 to 21.3) |
| Residual symptoms after discharge, n (%) |  |  |  |
| Chest congestion | 23 (39.7) | 170 (19.3) | 20.3 (7.1 to 33.6) |
| Cough | 15 (25.9) | 109 (12.4) | 13.5 (1.7 to 25.3) |
| Diarrhea | 5 (8.6) | 29 (3.3) | 5.3 (-2.2 to 12.9) |
| Dry eyes | 13 (22.4) | 96 (10.9) | 11.5 (0.3 to 22.7) |
| Expectoration | 11 (19.0) | 117 (13.3) | 5.7 (-5.0 to 16.3) |
| Fatigue | 11 (19.0) | 100 (11.4) | 7.6 (-3.0 to 18.2) |
| Fever | 5 (8.6) | 11 (1.3) | 7.4 (-0.1 to 14.8) |
| Headache | 5 (8.6) | 48 (5.5) | 3.2 (-4.4 to 10.8) |
| Myalgia | 6 (10.3) | 66 (7.5) | 4.1 (-5.4 to 11.1) |
| Palpitation | 13 (22.4) | 141 (16.0) | 6.4 (-4.9 to 17.8) |
| Sore throat | 6 (10.3) | 55 (6.3) | 4.1 (-4.1 to 12.3) |

For mean age, the difference is given in years; for mean length of hospital stay, the difference is given in days; for other characteristics, the difference is given in percentage points.

**Supplementary Table 4.** Age- and Sex-Adjusted Linear Regression Analysis for 11 Residual

**a) Chest congestion**
| **Variable** | **Coefficients (95% CI)** | **Coefficients Std. Error** | **p Value** |
| --- | --- | --- | --- |
| Age | 0.021 (0.011 to 0.032) | 0.005 | < 0.001 |
| Age^2 | 0.000 (0.000 to 0.000) | 0.000 | < 0.001 |
| Male | -0.024 (-0.076 to 0.027) | 0.026 | 0.350 |
| Re-positive test | 0.197 (0.090 to 0.303) | 0.054 | < 0.001 *** |

**b) Cough**
| **Variable** | **Coefficients (95% CI)** | **Coefficients Std. Error** | **p Value** |
| --- | --- | --- | --- |
| Age | 0.002 (-0.007 to 0.011) | 0.005 | 0.712 |
| Age^2 | 0.000 (0.000 to 0.000) | 0.000 | 0.762 |
| Male | -0.017 (-0.060 to 0.027) | 0.022 | 0.454 |
| Re-positive test | 0.135 (0.045 to 0.224) | 0.046 | 0.003 ** |

**c) Diarrhea**
| **Variable** | **Coefficients (95% CI)** | **Coefficients Std. Error** | **p Value** |
| --- | --- | --- | --- |
| Age | 0.003 (-0.002 to 0.008) | 0.003 | 0.242 |
| Age^2 | 0.000 (0.000 to 0.000) | 0.000 | 0.133 |
| Male | -0.004 (-0.028 to 0.020) | 0.012 | 0.760 |
| Re-positive test | 0.053 (0.003 to 0.102) | 0.025 | 0.037 * |

**d) Dry eyes**
| **Variable** | **Coefficients (95% CI)** | **Coefficients Std. Error** | **p Value** |
| --- | --- | --- | --- |
| Age | 0.013 (0.005 to 0.021) | 0.004 | 0.003 |
| Age^2 | 0.000 (0.000 to 0.000) | 0.000 | 0.001 |
| Male | 0.000 (-0.041 to 0.041) | 0.021 | 0.984 |
| Re-positive test | 0.250 (0.135 to 0.364) | 0.058 | 0.010 * |

**e) Expectoration**
| **Variable** | **Coefficients (95% CI)** | **Coefficients Std. Error** | **p Value** |
| --- | --- | --- | --- |
| Age | 0.008 (-0.002 to 0.017) | 0.005 | 0.103 |
| Age^2 | 0.000 (0.000 to 0.000) | 0.000 | 0.059 |
| Male | -0.005 (-0.049 to 0.039) | 0.023 | 0.833 |
| Re-positive test | 0.055 (-0.036 to 0.146) | 0.047 | 0.237 |

**f) Fatigue**
| Variable | Coefficients (95% CI) | Coefficients Std. Error | p Value |
| --- | --- | --- | --- |
| Age | 0.009 (0.000 to 0.017) | 0.004 | 0.041 |
| Age^2 | 0.000 (0.000 to 0.000) | 0.000 | 0.098 |
| Male | -0.035 (-0.077 to 0.006) | 0.021 | 0.096 |
| Re-positive test | 0.073 (-0.012 to 0.159) | 0.044 | 0.094 |

g) Fever
| Variable | Coefficients (95% CI) | Coefficients Std. Error | p Value |
| --- | --- | --- | --- |
| Age | 0.001 (-0.003 to 0.004) | 0.002 | 0.754 |
| Age^2 | 0.000 (0.000 to 0.000) | 0.000 | 0.690 |
| Male | 0.002 (-0.014 to -0.019) | 0.008 | 0.785 |
| Re-positive test | 0.074 (0.039 to 0.108) | 0.017 | < 0.001 *** |

h) Headache
| Variable | Coefficients (95% CI) | Coefficients Std. Error | p Value |
| --- | --- | --- | --- |
| Age | 0.005 (-0.001 to 0.011) | 0.003 | 0.096 |
| Age^2 | 0.000 (0.000 to 0.000) | 0.000 | 0.104 |
| Male | -0.007 (-0.036 to 0.023) | 0.015 | 0.659 |
| Re-positive test | 0.030 (-0.031 to 0.092) | 0.031 | 0.337 |

i) Myalgia
| Variable | Coefficients (95% CI) | Coefficients Std. Error | p Value |
| --- | --- | --- | --- |
| Age | 0.008 (0.001 to 0.015) | 0.004 | 0.022 |
| Age^2 | 0.000 (0.000 to 0.000) | 0.000 | 0.020 |
| Male | -0.019 (-0.053 to 0.015) | 0.017 | 0.274 |
| Re-positive test | 0.026 (-0.044 to 0.097) | 0.036 | 0.464 |

j) Palpitation
| Variable | Coefficients (95% CI) | Coefficients Std. Error | p Value |
| --- | --- | --- | --- |
| Age | 0.012 (0.002 to 0.021) | 0.005 | 0.019 |
| Age^2 | 0.000 (0.000 to 0.000) | 0.000 | 0.018 |
| Male | 0.004 (-0.044 to 0.051) | 0.024 | 0.879 |
| Re-positive test | 0.060 (-0.038 to 0.159) | 0.050 | 0.232 |

k) Sore throat
| Variable | Coefficients (95% CI) | Coefficients Std. Error | p Value |
| --- | --- | --- | --- |
| Age | 0.003 (-0.003 to 0.010) | 0.003 | 0.300 |
| Age^2 | 0.000 (0.000 to 0.000) | 0.000 | 0.172 |
| Male | -0.014 (-0.046 to 0.018) | 0.016 | 0.391 |
| Re-positive test | 0.041 (-0.025 to 0.106) | 0.033 | 0.222 |

\* p < 0.05 \*\*p < 0.01 \*\*\*p < 0.001

**Supplementary Table 5.** Coexisting Diseases of RP and RN patients above 50

| Coexisting diseases, n (%) | RP Patients<br>(n=43) | RN Patients<br>(n=638) | Difference<br>(95% CI) |
| --- | --- | --- | --- |
| Diabetes mellitus | 10 (23.2) | 103 (16.1) | 7.1 (-6.3 to 20.6) |
| Coronary artery disease | 9 (20.9) | 55 (8.6) | 12.3 (-0.5 to 25.1) |
| Hypertension | 20 (46.5) | 190 (29.8) | 16.7 (0.8 to 32.6) |
| Diabetes mellitus and coronary artery disease | 6 (14.0) | 12 (1.9) | 12.1 (1.2 to 22.9) |
| Coronary artery disease and hypertension | 7 (16.3) | 28 (4.4) | 11.9 (0.3 to 23.5) |
| Diabetes mellitus and hypertension | 8 (18.6) | 49 (7.7) | 10.9 (-1.4 to 23.2) |

The difference is given in percentage points.

**Supplementary Table 6.** Age- and Sex-Adjusted Linear Regression Analysis for RP Patients over 50

a) Diabetes mellitus
| Variable | Coefficients<br>(95% CI) | Coefficients Std.<br>Error | p Value |
| --- | --- | --- | --- |
| Age | 0.019 (-0.007 to 0.045) | 0.013 | 0.151 |
| Age^2 | 0.000 (0.000 to 0.000) | 0.000 | 0.137 |
| Male | 0.005 (-0.044 to 0.054) | 0.025 | 0.843 |
| Diabetes mellitus | 0.032 (-0.017 to 0.081) | 0.025 | 0.202 |

b) Coronary artery disease
| Variable | Coefficients<br>(95% CI) | Coefficients Std.<br>Error | p Value |
| --- | --- | --- | --- |
| Age | 0.020 (-0.006 to 0.045) | 0.013 | 0.134 |
| Age^2 | 0.000 (0.000 to 0.000) | 0.000 | 0.117 |
| Male | 0.006 (-0.042 to 0.054) | 0.025 | 0.811 |
| Coronary artery disease | 0.089 (0.026 to 0.151) | 0.032 | 0.006 ** |

c) Hypertension
| Variable | Coefficients<br>(95% CI) | Coefficients Std.<br>Error | p Value |
| --- | --- | --- | --- |
| Age | 0.018 (-0.007 to 0.044) | 0.013 | 0.161 |
| Age^2 | 0.000 (0.000 to 0.000) | 0.000 | 0.141 |
| Male | 0.003 (-0.046 to 0.051) | 0.025 | 0.906 |
| Hypertension | 0.048 (0.008 to 0.088) | 0.020 | 0.018 * |

d) Diabetes mellitus and coronary artery disease
| Variable | Coefficients<br>(95% CI) | Coefficients Std.<br>Error | p Value |
| --- | --- | --- | --- |
| Age | 0.021 (-0.004 to 0.046) | 0.013 | 0.102 |
| Age^2 | 0.000 (0.000 to 0.000) | 0.000 | 0.086 |
| Male | 0.007 (-0.041 to 0.055) | 0.024 | 0.778 |
| Diabetes mellitus and coronary artery disease | 0.286 (0.173 to 0.398) | 0.057 | < 0.001 *** |

e) Coronary artery disease and hypertension
| Variable | Coefficients<br>(95% CI) | Coefficients Std.<br>Error | p Value |
| --- | --- | --- | --- |
| Age | 0.019 (-0.006 to 0.045) | 0.013 | 0.136 |
| Age^2 | 0.000 (0.000 to 0.000) | 0.000 | 0.122 |
| Male | 0.005 (-0.043 to 0.054) | 0.025 | 0.829 |
| Coronary artery disease and diabetes mellitus | 0.146 (0.064 to 0.229) | 0.042 | 0.001 ** |

f) Diabetes mellitus and hypertension
| Variable | Coefficients<br>(95% CI) | Coefficients Std.<br>Error | p Value |
| --- | --- | --- | --- |
| Age | 0.019 (-0.007 to 0.044) | 0.013 | 0.152 |
| Age^2 | 0.000 (0.000 to 0.000) | 0.000 | 0.133 |
| Male | 0.004 (-0.044 to 0.053) | 0.025 | 0.861 |
| Diabetes mellitus and hypertension | 0.087 (0.021 to 0.153) | 0.034 | 0.010 * |

\* p < 0.05 \*\* p < 0.01 \*\*\* p < 0.001

